# Virtually in Synch: A Pilot Study on Affective Dimensions of Dancing with Parkinson’s during COVID-19

**DOI:** 10.1101/2021.05.17.20249000

**Authors:** Katayoun Ghanai, Rebecca E. Barnstaple, Joseph FX DeSouza

## Abstract

Loss of social supports and community programs due to lockdowns and other measures associated with COVID-19 has been linked with concerns over mental health and feelings of isolation. These challenges can be particularly acute for the elderly and people living with chronic or pervasive health conditions. *Dance for PD*, a program specifically developed for people living with Parkinson’s Disease, formerly offered in hundreds of locations around the globe, either halted or shifted to a virtual format. Our study investigates the transition of these dance-based programs to an online environment, with the aim of determining the extent to which a virtual format provides affective support or other benefits. Given the increased incidence of mental health problems and social isolation associated with COVID-19, this investigation aims to contribute to the development of better supports for vulnerable populations while helping us understand the specific contributions of dance-based programs in a virtual environment.

## Introduction

COVID-19 has changed our world, and the increased uncertainty, fear, and isolation that have come in its wake are causing widespread concern for mental health. Loss of social supports and community has disproportionately affected the elderly, for whom separation from loved ones, and loss of access to programs providing opportunities for social connection has an even greater impact. The dance community has shown tremendous creativity in response to the constraints and challenges of COVID-19, including the extended community of dance educators who work with people diagnosed with Parkinson’s Disease (PD). Over the past 19 years, *Dance for PD* and affiliates have supported the development and delivery of programs around the world, many of which have now shifted to a virtual format. Our study investigates the impact of the transition to the online environment for participants in these programs, with the aim of determining the extent to which a virtual format can provide affective and other benefits to people living with PD. Given the increased incidence of mental health problems and isolation associated with COVID-19, this investigation aims to contribute to the development of better supports for this and other vulnerable populations.

Parkinson’s disease (PD) is a degenerative neurological disorder affecting more than 10 million people worldwide, most of whom are over the age of 55. Based on the statistics of the Parkinson’s Foundation, each year about 60,000 Americans are diagnosed with the disease with this number expected to be circa 1,000,000 in 2020 in the US alone. Although Parkinson’s primarily impacts older adults, 4% of people diagnosed in America are under the age of 50. The direct and indirect costs of Parkinson’s are estimated at $52 billion annually for the US. Annual medication costs for PD average USD 2500 per individual.^1^ The increasing incidence of PD has a significant effect on society, both in terms of professional care and the burden on private care givers (Dorsey et al., 2018). While PD itself is progressive and often debilitating, ensuring quality of life and standards of care are challenges that are distributed amongst care partners, medical professionals, and the person living with the disease. PD symptoms emerge when neurons in the basal ganglia (BG) responsible for the production of dopamine (specifically the substantia nigra pars compacta SNc) are reduced to a critical level, at which point the balance of dopamine is not sufficient to maintain function, leading to progressive and pervasive symptoms. Motor symptoms appear when more than ∼80% of dopamine producing cells in the SNc are lost, indicating that the onset of PD likely occurs years before the appearance of symptoms. Motor symptoms include tremors, bradykinesia, gait and balance problems, and limb rigidity. Non-motor symptoms that are notable prior to the appearance of motor symptoms include loss of olfaction, mood changes, anxiety, and REM-sleep behaviour disorder (Jankovic, 2008). The cause of damage to dopaminergic neurons is not yet known, and while current pharmacological treatments (notably L-dopa) can decrease the impact of symptoms, their efficacy decays over time, and high dosages can cause side effects impacting breathing, mood, and movement including dystonia. Extrapyramidal side effects (EPS) which resemble dyskinesia or motor dysfunctions can also occur, attributable to the decay of the medication rather than the disease itself.

While PD is usually considered in terms of its motor symptoms, negative affective elements that significantly impact quality of life are also common. Depression is considered prodromal to the development of PD (Pellicano et al., 2007), and approximately 40% of people with PD are also diagnosed with depression (Cummings, 1992). Abnormal activations of reward circuitry in the brain have been implicated in major depressive disorder (Ng, Alloy & Smith, 2019) and impulse control disorder (ICD) in PD has been shown to be associated with reward hypersensitivity (Drew et al, 2020). The synchronisation of movements with music that happens in dance, and the process of acquiring skilfulness in learning and performing movement, activates this circuit (Bar & DeSouza, 2016). The reward network described by Berridge & Kringelbach (2015) contains anatomical regions of prefrontal cortex (PFC), including portions of orbitofrontal, insula, and anterior cingulate cortices (ACC), as well as subcortical limbic structures such as the NAc, ventral pallidum (VP), and amygdala. These regions also monitor where the body is in space via the insula circuitry, another crucial skill that is trained and developed through participation in dance. The orbitofrontal cortex, a key node in the network, is an important nexus for sensory integration, emotional and hedonic processing. The reward center is located in mesolimbic dopamine pathway which connects the nucleus accumbens (NAc) with the ventral tegmental area (VTA) where dopamine is also produced (Alberico, 2015). VTA neurons have been shown to degenerate in PD (Alberico, 2015) and this degeneration is associated with nonmotor symptoms such as depression. Participating in dance has been shown to have a positive effect on depression (Akandere et al., 2011; Koch et al., 2007; Ciantar et al., 2019), suggesting that it may contribute to reducing the likelihood or incidence of depression in people with PD who are deficient in dopamine. To investigate this hypothesis, we conducted a pilot study to determine whether online dance programs would also have a positive role in elevating the mood of people with PD, comparable to affective changes recently shown in live classes (Fontanesi & DeSouza, 2020). This would demonstrate that *Dance for PD* classes continue to be effective in modulating mood in an online environment.

## Dance with PD

While PD has no known cure and few treatment options, there is a growing body of research demonstrating a role for dance in helping people with this diagnosis. Dance-based programs have been shown to reduce motor and non-motor symptoms associated with PD (Bearss et al., 2017; Hackney et al., 2007; Heiberger et al., 2011, Westheimer et al., 2008; Westheimer et al., 2015), diminish negative affective symptoms (Ciantar et al., 2019; Fontanesi & DeSouza, 2021), improve quality of life (Bearss et al., 2017; Heiberger et al., 2011; Westheimer et al., 2008; Westheimer et al., 2015), and even potentially slow disease progression over 3 years (DeSouza & Bearss, 2018).

Participation in dance and learning new movement strategies can support simultaneous improvements in motor and cognitive domains (Hashimoto et al., 2015), and Bognar et al. (2017) have shown that dance has a positive effect on the social self, concluding that participation in dance may help people with PD cope with emotional challenges. Our study assesses the impact of COVID-19 related transitions on the efficacy of the largest and most established dance-based method for people living with PD: *Dance for PD*, founded in 2001 by the Mark Morris Dance Group in Brooklyn, New York, includes a network of affiliates in more than 200 communities in 25 countries around the world (www.danceforparkinsons.org/about-the-program).

## Structure of Live dance classes with PD before COVID-19 Pandemic

Pre-pandemic, most *Dance for PD* classes were an hour in duration, once a week, with a certified dance instructor and live music wherever available (sites following this format include National Ballet of Canada and Trinity in Toronto). Each class had a number of volunteers who participate in class and assist when needed.

Dance elements of class include combinations of jazz steps, ballet, Argentinian tango, dance theatre, free style and choreographed movements.

Based on Mark Morris technique, most classes followed the routine below:

1. Welcome/Opening; introductory game such as a name game.
2. Warming up while seated and breathing exercises.
3. Movements for upper body while seated.
4. Movements for lower body while seated.
5. Paired dance while seated, may include mirroring.
6. Standing sequences close to the chair, such as *pliés*.
7. Locomotion through the room.
8. Free style or improvisatory movement.
9. Closing; in a circle, holding hands, and bowing to each other.
10. A special thank you to the piano player from everybody.

Each portion of the class would be accompanied by a specific rhythm, the music matched with the narrative provided by the choreographer/teacher. For example, the instructor may choose a theme of swimming in a beautiful ocean under the sun for a cold winter day. The teacher would describe the scene or imagery, then model or explain the movements. When live accompaniment was available, the musician would either improvise or play a theme arranged to fit the story line. The instructor would ask the musician to increase or decrease the tempo based on the mood or capability of the dancers in each session. The musician would continue playing as long as the instructor continued the narration.

While this model remains congruent across sites and *Dance for PD* teachers receive the same training, there is room to develop a unique class flavour based on the teacher’s background, local culture, and the interests of the dancers. In Toronto, classes offered by *Dancing with* Parkinson’s at Trinity were often more theatrical, with extensive room for dancers to exercise their imaginations and creativity. Participants formed a strong sense of community over their years and months of dancing together and looked forward to attending class. Even when people were not feeling optimal, they would make an effort to attend as they knew they would feel better afterwards. At National Ballet School (NBS) participants often spent time together after class in the atrium to talk and share cookies and coffee. Opportunities for socialising built into the *Dance for PD* model, are another aspect for this program that contributes to enhancing quality of life for participants. Heiberger et al. (2011) studied the effects of dance more specifically on quality of life for people with PD using the Quality-of-Life Scale (QOLS) and Westheimer questionnaire, and found significant positive effects. Research in our own group of new dancers who started in September 2013 confirmed these effect (Bears et al, 2017).

## Structure of On-Line Dance Classes

Online dance classes have shown more variety in format, with frequencies ranging from once a week to daily, and class lengths of 20 minutes to 1+hour in length. *Dancing with Parkinson’s* in Toronto, Canada is running classes every day at 11:00 EST for ∼20 minutes via Zoom. These are entirely seated and follow a similar routine to the seated portion of previously live classes. Musical accompaniment is recorded and often has lyrics. Each day, approximately 60 people on average attend. As the class is held online and anyone with internet in their home can easily access Zoom, elderly participants without Parkinson’s have also been given the opportunity to join. Most people join with their cameras off, and the instructor is in “spotlight”. Many people in this group did not know each other previously, and although there is some talking at the end of the class, they don’t have the opportunity to connect with each other socially. Most conversation is about the quality of internet connection, sound issues, or questions regarding the movement material. The instructor is also limited by these conditions in their ability to connect with the dancers and respond through music or movement material. Sound quality may be affected by the need to speak and demonstrate movement at the same time, while the music is also broadcast over the same microphone. There has yet to be a fundamental review of different approaches to better understand factors making online classes more accessible. For best practices to emerge in the design and implementation of practical long-term virtual programming, data that can inform dance-based programs involving people living with PD is of particular importance.

## Background

Our group has been conducting research on the effects of dance for PD since 2013, and data collected thus far include multimodal neuroimaging (fMRI, EEG; Harrar et al 2016; Levkov et al., 2014), mood (GDS; PANAS-X; Ciantar et al., 2019; Fontanesi & DeSouza. 2021), and motor performance (Berg, TUG, UPDRS; Bearss et al., 2017; Bearss & DeSouza, 2017; DeSouza & Bearss, 2018). Participants have also completed detailed questionnaires concerning medication, motor, and non-motor problems to separate EPS from PD symptoms. PANAS-X is a self-reported questionnaire referring to the Positive/ Negative Affect Schedule, measuring higher and lower order emotional states representative of mood. Based on Watson and Clark, PANAS-X divides emotions into three categories: basic negative emotional states such as fear and sadness, basic positive emotional states such as cheerfulness and daring, and other affective states such as inspired and interested (Watson & Clark, 1999). Preliminary results in previously collected in-person data showed positive improvements in mood related symptoms for people with PD who participated in dance (Ciantar et al., 2019). We have also recorded improvements in motor symptoms from TUG, BERG and UPDRS (Bearss et al., 2017; Bearss & DeSouza, 2017; DeSouza & Bearss, 2018), in line with other studies that have shown decreases in motor symptom measures such as TUG or functional mobility (Dos Santos Delabary et al., 2018). Through neuroimaging studies using fMRI, we have preliminary evidence of brain-related changes associated with dance in expert dancers (DeSouza & Bar, 2012; Bar and DeSouza, 2016) and people with PD (Harrar et al., 2016) and changes in neural signalling patterns through rsEEG (Bearss & DeSouza, 2017; Levkov et al., 2014).

Previous research has focussed on the impact of “live” classes held in studio with one or more teachers, volunteers, and students all attending in-person; COVID-19, however, forced the closure of *Dance for PD* classes around the world. While many *Dance for PD* classes have retained, or attempted to replicate, the style of class that was offered prior to COVID-19, shifting online has been accompanied by structural changes for some organizers. These include changes in the length of class time (in some cases, a 75-minute class has been shortened to 20 minutes), frequency (in Toronto, classes are now offered daily for all participants), less inclusion of standing/supported standing, and less movement through space. For the current study, we hypothesized that dancing online would have a positive role on Affect, elevating the mood of people with PD, comparable to that recently shown in live classes (Fontanesi & DeSouza, 2021). This would demonstrate that moving to music in this way can be powerful and emotionally potent for people with PD, even with the changes that are necessitated by an online environment.

## Methods

Following ethics approval for a study involving human participants from Office of Research Ethics at York University with the approval of all questionnaires and procedures (Certificate# 2017-269), an online survey including a consent form and PANAS-X evaluating mood before and after the virtual class, along with questions pertaining to attendance, music type, and enjoyment, were distributed through an on-line Qualtrics link to classes around the world. As the population is elderly and may be less familiar with technology, data collection can require longer to complete, and responses are currently ongoing. Participants in this report include 17 females and 5 males with a mean age of 68, diagnosed with Parkinson’s disease. Participants were requested to fill out a consent form and Pre PANAS-X before attending their online dance class, and to note the time at which they finished the Pre and at which they commenced the Post, in order for us to ensure the accuracy of Post PANAS-X which needed to be completed immediately after class.

Requests for participation were sent to 38 organizers of *Dance for PD* classes around the world. Some organizers did not respond to our request. 20 teachers or organizers announced their interest in our study and the numbers they reported are shown in Figure 2. Amongst the 20 respondents, only 15 were conducting online classes. Four organizers replied that they did not feel they could ask their dancers to participate in our study due to Corona fatigue or another study having been recently conducted with their group. The groups who showed interest in participating shared the online Qualtrics link with their participants during their online dance classes. We received a total of 51 responses to our online questionnaire, of which 30 questionnaires were not fully completed; this could be due to participants forgetting to fill out the second part of the PANAS-X after class or becoming fatigued and leaving their computers. Analytical statistics were completed for the remaining 21 completely filled questionnaires. Among these, four people did not complete the Post PANAS-X part of the survey and could not be included in the paired t-test evaluation of mood. Data analysis was performed by three lab members who were not engaged in this research topic.

**Figure 1.**
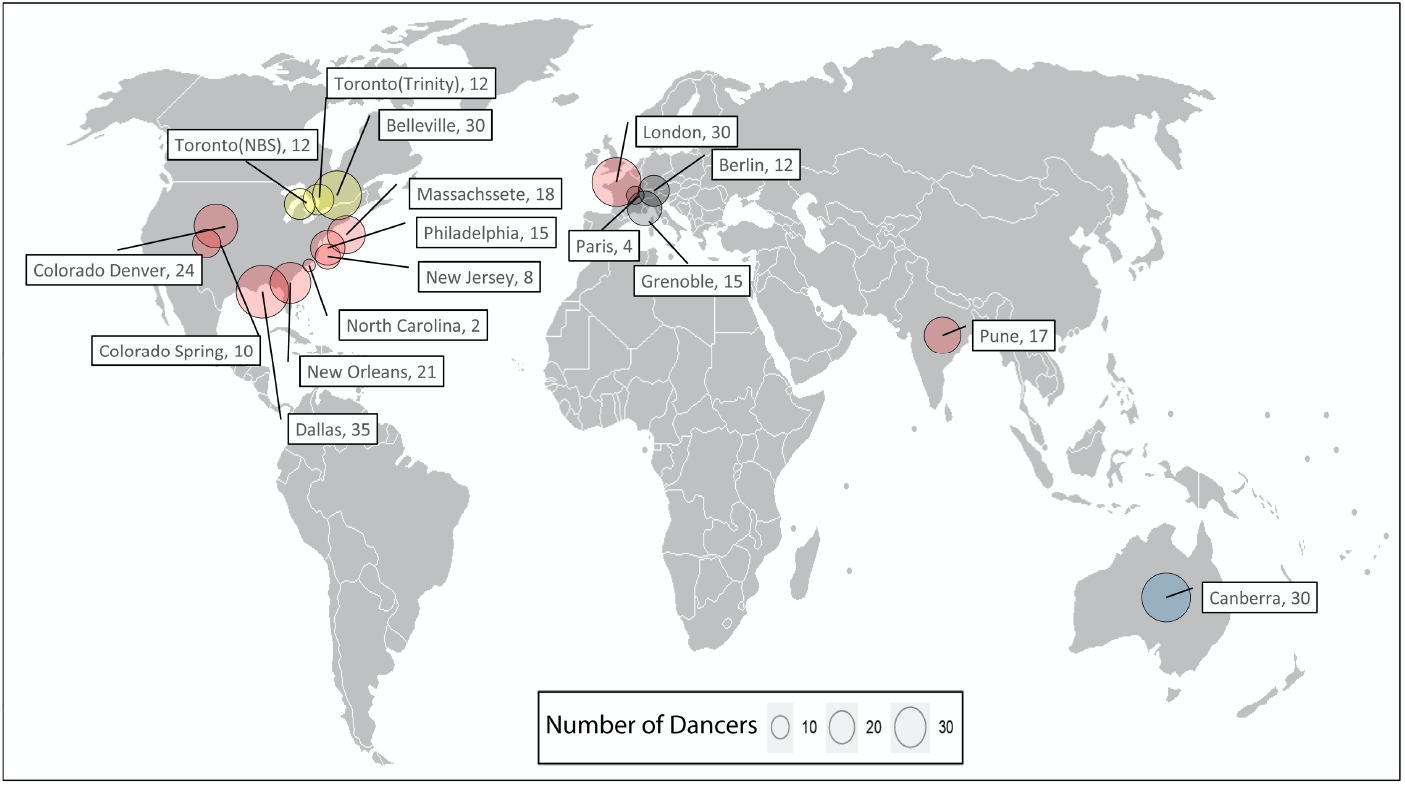
World map showing the average number of people with PD who were dancing in participating studios before the pandemic based on our survey from November 30, 2020.

**Figure 2.**
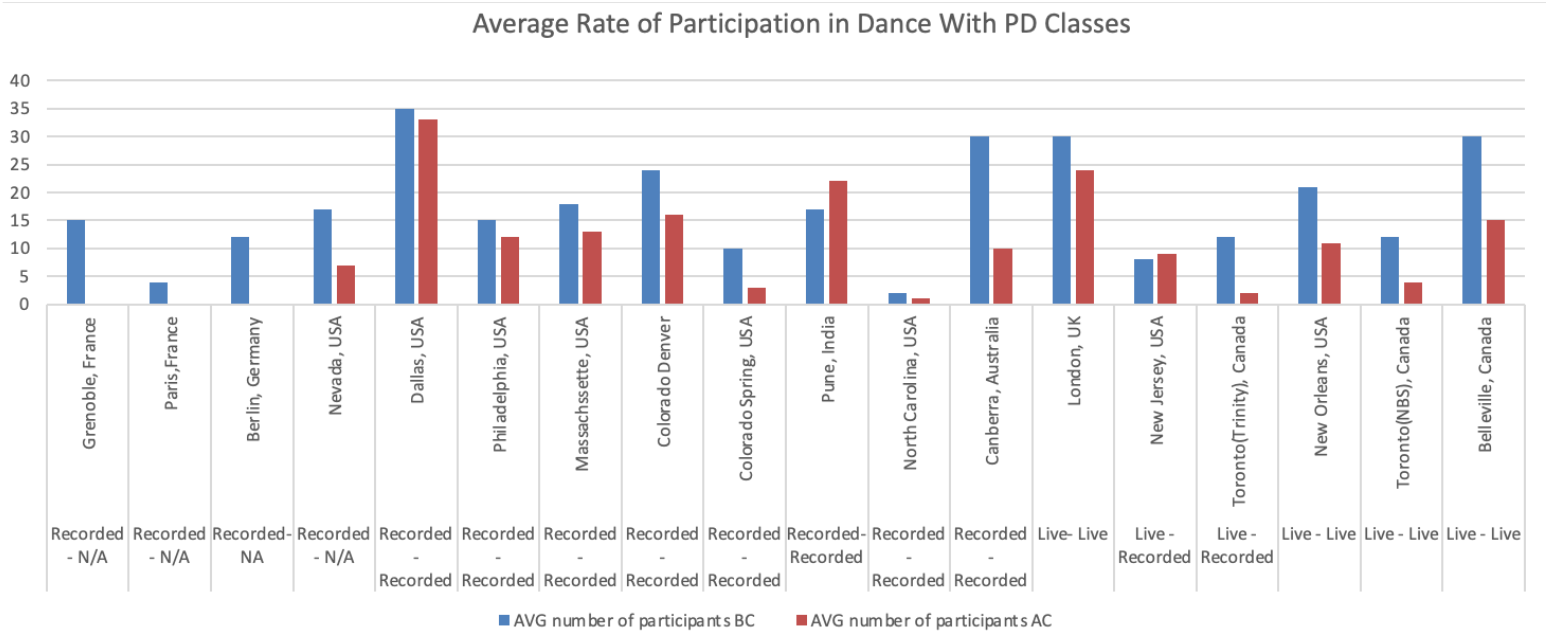
Bar chart showing the average number of participants of DWP before Corona-19. (BC) and after Corona-19 (AC) in different countries with the music used in classes BC and AC.

## Results

From the responses of the organizers who responded to our survey, on average 281 people with PD were dancing in participating studios before the Pandemic (Figure 1). After COVID-19 pushed classes online, an average of 182 people with PD reported attending virtual classes at these locations (Figure 2), representing more than 64% of the population for our study of dancers with PD. It is common in organized sessions for PD that the number of participants will fluctuate; prior to COVID-19 participation in this type of class is a daily decision based on an individual’s current physical condition. Thus, we used averages of the minimum and maximum number of the participants in each class for our study.

Most importantly, when examining the results of the potential affective changes before and after a single online dance class, we performed a pre/post-dance scoring for two general dimensions of affect: General Negative Affect (GNA) and General Positive Affect (GPA). Figure 3 shows the GNA and GPA for all 17 participants. The first set of bars represent a significant reduction in the median Negative affect before (orange bars) and after class (green bars; paired *t*-test, t= 2.9059, df=16, *p*=0.01031, alternative hypothesis: true difference in means is not equal to 0.95 percent confidence interval: 0.8910064 - 5.6972289). Similarly, the positive affect median scores increased after class (paired *t*-test, t=-3.01833, df= 16, *p*=0.005777). The collapsed negative and positive affect sum score also showed a significant difference, t=-3.1833, df= 16, *p*=0.005777. Effect size: d estimate: -0.4440409. 95 percent confidence interval: lower -0.7418541. upper -0.1462277. All comparisons are significant at the Bonferroni corrected alpha significance level at *p* < (0.05 / 3) = *pCorr* < 0.017. The interpretation of the data analysis based an alternative hypothesis (there is a difference in the mood of the dancers in virtual classes before and after dancing) confirms a significant difference in the mood of the dancers before and after their online dance class (t-value= -3.0183, P<0.001).

**Figure 3.**
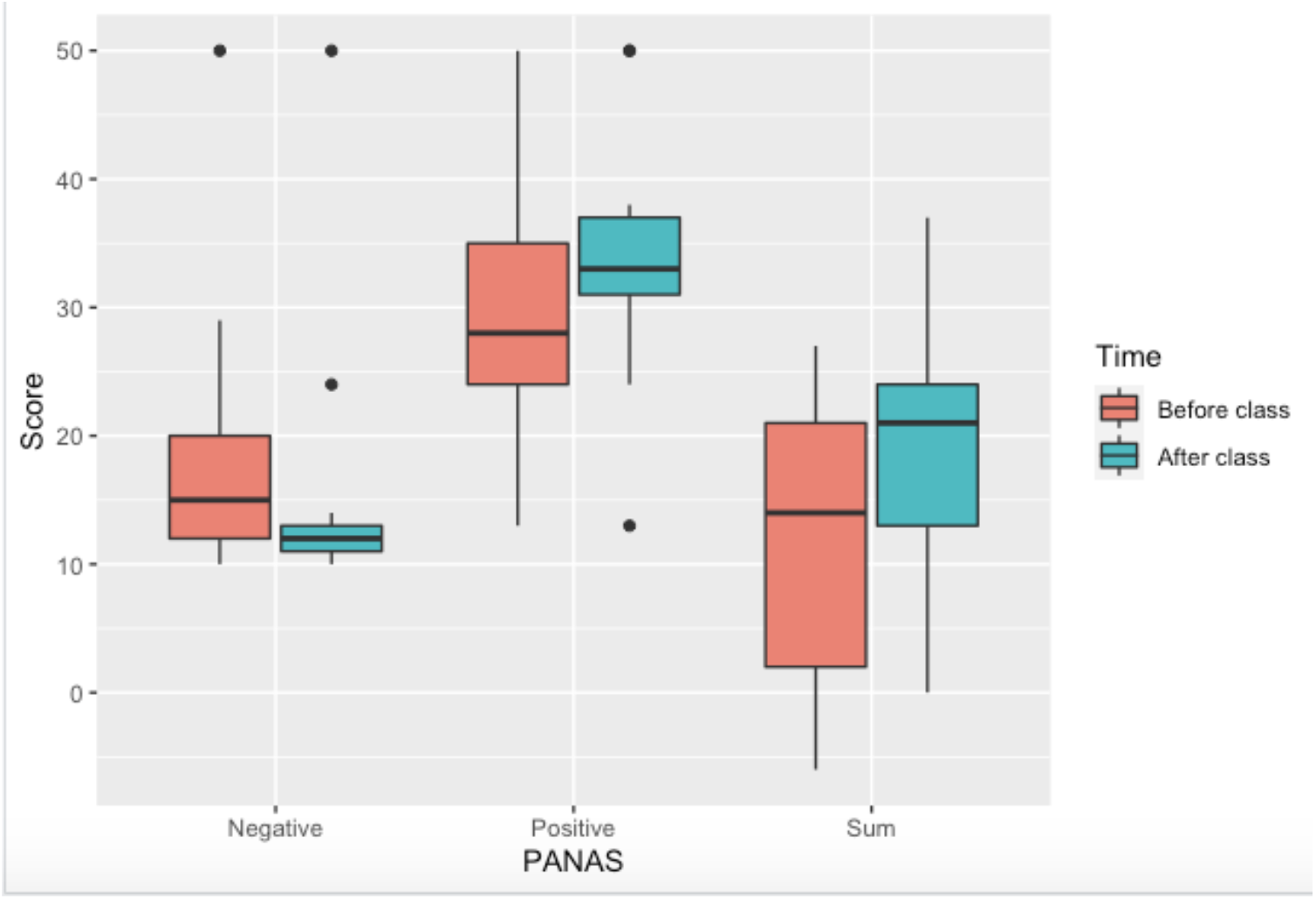
Results from PANAS-X before and after 1 online dance class: Negative Affect after dancing online decreased, Positive Affect after dancing online increased, and the sum of Positive Affect minus Negative Affect shows an improvement in mood after a single online class. The boxes show from 25% to 75% of the scores; the line in the middle is the median. The lines above and below the boxes represent the remainder of scores; dots are outliers.

In addition to these significant affective improvements for people participating in dance classes online, a marked preference for live classes was expressed in our questionnaire, associated with appreciation of social interactions that happen in live classes (Figure 4). Reasons for this preference, based on our survey data, were primarily issues such as difficulty with using the technology and/or a preference for live music; however, missing social interactions contributed a significantly higher index to the preference for live over virtual formats. 13 people out of 21 answered that they preferred live classes because of social interactions. Respondents who preferred online over live classes said this was due to transportation issues (three people) or a preference for recorded music (one person); this portion of responses scarcely compares with the number of people who preferred live classes and missed social engagement (13 people). Two people had no preference between live or virtual classes.

**Figure 4.**
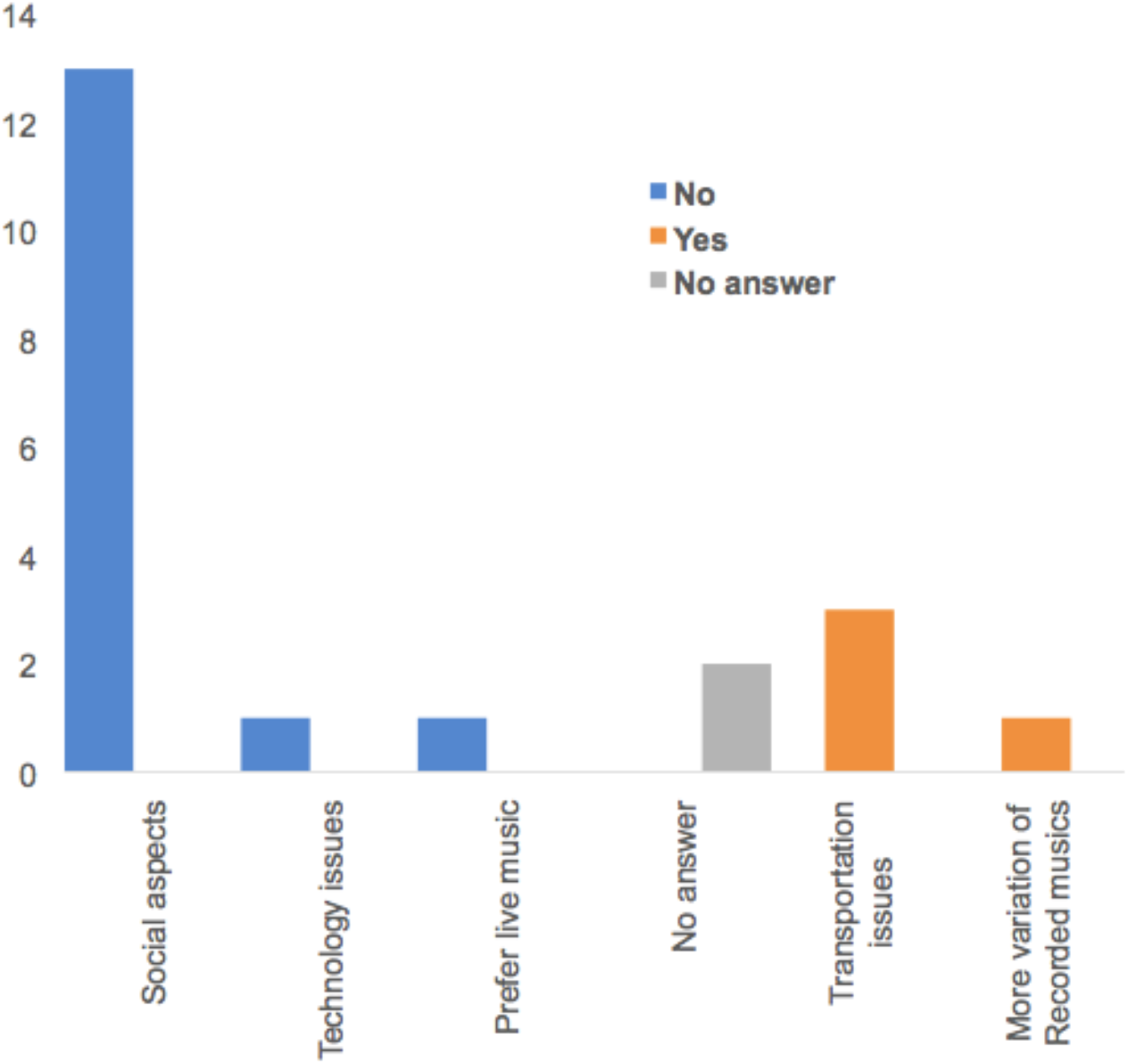
Question: Do you prefer online classes to live ones? Why? Most people who did not prefer dancing online miss the social aspects of live classes.

## Discussion

According to choreographer Mark Dendy, “Dance is a contemporary ritual and a spiritual opportunity to share energy, join in a sense of *communitas*” (Dils et al., 2001). The newly established UNESCO Chair on Dance and Social Inclusion (University of Auckland) has been created in recognition of the role that dance can play in building community and recentering excluded bodies in global discourse. Health and wellbeing are an integral part of the type of inclusion that can be built by and through participation in dance. Receiving a diagnosis of a neurodegenerative disease such as Parkinson’s and losing capacity for movement and agency during a global pandemic can be a frightening experience. The shift to online delivery formats for essential arts-based programs and services such as *Dance for PD* has played an important role in buoying the spirits and hopes of people living with PD and their care partners. Having the opportunity to continue a familiar movement practice, or try it for the first time, in a supportive atmosphere within an online community clearly impacts mood and wellbeing.

People living with PD, and the dance educators who work with them, have shown incredible resilience in their ability to adapt to new circumstances. The results of our study demonstrate that virtual programs offer important affective support for people living with PD. This diagnosis can lead to social isolation due to impediments associated with the disease, and dance for PD programs provide an opportunity to build community among others with a shared experience. Worldwide mandatory isolation orders and virtualization of community programs and supports due to COVID-19 have led to heavy burdens, both financial and emotional, for people with PD and their caregivers. The level of participation from members of the PD community may be an incentive for the organizers to focus their research on strategies that motivate people with PD to join online classes. Participation may be aided by the attitudes of organizers and further research demonstrating the efficacy of programs such as *Dance for PD*.

Reports from organizers that some seniors find the online format better meets their needs than in-person classes suggest that there will likely be a demand for virtual dance into the future. Some locations have even reported an increase in participation; pre-COVID, New York had nine locations offering *Dance for PD* classes. After the transition to virtual, the number of people who signed up for online classes with Mark Morris group in New York increased significantly; based on an interview with program director David Leventhal in Dance magazine September 2020, the number of participants more than doubled, from 600 to 1500. Leventhal mentions that 10% of the new participants are from New York who probably had transportation issues for in person classes. He suggests that online attendance offers participants some degree of privacy while partaking in the benefits dance; people who don’t wish to share their PD diagnosis or who are timid about their appearance can attend with cameras off. Another benefit of online classes in Leventhal’s view is the possibility to access to classes in other languages. Mark Morris group is launching classes in Spanish with collaboration of Muhammad Ali Parkinson’s center in Phoenix Arizona in 2021 (from https://www.dancemagazine.com/dance-for-pd2647781883.html?rebelltitem=2#rebelltitem2). Sarah Robichaud, a teacher and organizer from Toronto, Ontario has been offering classes seven days a week since the start of the pandemic through her organisation *Dancing with Parkinson’s* (DWP), and participant numbers continue to grow. She attributes this to the convenience in accessing classes online, particularly for seniors with PD for whom organizing transportation and moving through the city may be difficult, and says participants in her programs find there can be a greater degree of intimacy and less distraction in participating from home (https://torontoevaluation.ca/blog/index.php/2020/05/14/adapting-a-dance-intervention-in-the-time-of-covid-19/). While remaining at home may be both convenient and prudent at this time, it is essential that we all continue to challenge ourselves and find opportunities for movement and learning. As class organizer Hrishikesh Pawar In Pune, India stated in his interview with us, “the more people stay at home, the more they need to move.”

Previous research conducted by our group on in-person *Dance for PD* classes showed that while depressive symptoms improved after attending class (as measured by the Geriatric Depression Scale/GDS), they dramatically increased during the three-month period over which regular weekly dance classes were not offered due to summer holidays (Ciantar et al., 2019). This suggests that virtual classes answer to an important need during COVID-19, and while they may not offer exactly the same benefits as “live” classes, they are infinitely better than no classes at all. A recent paper by Quinn and colleagues (2020) followed 27 patients with PD over two months following the onset of COVID-19 and concluded that virtual classes are a sustainable platform for coaching and maintaining the health of people with PD who can’t easily access live classes. Addressing motor symptoms, including gait and balance, together with supports for depression, should be considered in the type of physical exercise involved (Quinn et al., 2020). Dance, which offers integrated motor-cognitive training and learning opportunities, follows this model by promoting movement in a complex, artistic, and socially rich environment.

The relationship between exercise type and depressive symptoms can be assessed through mood evaluation with PANAS-X, as performed in our brief study on online dancing post-COVID in the PD community. Although our sample size is small, it is comparable to other dance studies, and the t-test is significant in showing important affective changes immediately after dancing online. It is important that we consider and note other elements that may be involved in elevating mood in addition to dance and music. The lack of social engagement and close contact with loved ones, friends, and community members during COVID-19 negatively impacts mental health (https://www.kff.org/coronavirus-covid-19/issue-brief/the-implications-of-covid-19-for-mental-health-and-substance-use/). People living with a progressive diagnosis such as PD which also affects mobility can already have a limited interaction with the outside world; attending classes online and feeling part of a virtual community may offer some relief for feelings of isolation during this pandemic.

Dance clearly has an impact on affect for people with PD but having access to a studio setting plays an important role, when available, in supporting social interaction. As previously mentioned, most people who participated in our survey were dancing in studios before the pandemic. However, many other factors likely influence the continued participation of past participants or the addition of new ones, including socio-economic status, urban versus rural settings, the availability of other resources and programs, access to technology and the internet, and computer literacy. Further research is needed to better understand the influence of these factors and other elements involved in complex, holistic, arts-based interventions such as *Dance for PD*. Future studies on the potential for programs such as *Dance for PD* and other hybrid music and dance classes to impact multidimensional aspects of mood and function will also greatly benefit from interdisciplinary approaches bringing together patients, clinicians, researchers, and program providers. Emphasis on discovering and promoting accessible ways of improving quality of life for people with PD, such as online classes, will be beneficial in managing neurodegenerative diseases in future waves of COVID-19 and beyond.

## Limitations and Future research

This report is based on a sample of people dancing during COVID-19 from the period inclusive of October 22^nd^ to Nov 30^th^ of 2020. Our study involves the relatively small population sample that completed all aspects of our survey; however, the power is comparable to other dance studies. Even within this sample we saw significant affective change associated with a single virtual dance class. We will continue collecting data for future follow-up studies examining future waves of the COVID-19 Pandemic. Future questions of importance could include details as to the duration of effects - how long is enhanced mood experienced after class? We have an opportunity during the pandemic to investigate through further at-home assessment tools what, if any, effects are felt 5-hrs after dance class, before bed, or the next morning, to examine the wave of mood modulation in a behavioral pattern online. From previous research (Ciantar et al. 2019) our group has observed a decay in mood effects during a 3-month period in which participants had no access to dance. Samples taken now, during the pandemic, should also be resampled to assess mood effects across time (days, weeks, months) once in-person classes start again.

Online survey formats can be somewhat challenging for older participants, and this may explain why a percentage of responses were incomplete. Language barriers may have also contributed to this issue. Our research included many countries and people with PD who may not have been fluent in English; however, as Dance for PD teachers are trained by the Mark Morris foundation in New York and are thus comfortable with English, we had good responses from teachers. Fatigue may have also contributed to facilitators and potential participants declining to be involved in our study. Many respondents who did not wish to participate said they had been involved in other projects and could not be asked to participate in another study at this time. Future iterations of our study may use a shorter version of PANAS-X, translated into other languages, and we will continue to research and develop user-friendly assessment tools that facilitate and improve responses.

Our study is one of the first to investigate the affective impact of online dance programs for people with PD during the Pandemic. This is a rich and complex topic that can be studied from many angles to better understand key components contributing to efficacy and may lead to the development and proliferation of optimal programs for people with PD, which have applications for other populations, particularly those experiencing challenges related to aging and isolation. Particularly during this time of lockdowns and closure of facilities, exercise at home can help people with PD and potentially enhance an immune response against COVID-19 symptoms (Hall et al., 2020). Our sample population was small relative to the average number of people who are participating in virtual classes all around the world. Further research can be conducted with the collaboration of more classes around the world and sharing ideas between researchers, program providers, and participants.

## Data Availability

Yes data will be available from our lab website

http://www.joelab.com/katyDataset.zip

## Acknowledgments

The authors declare that the research was conducted in the absence of any commercial or financial relationships that could be construed as a potential conflict of interest.

## Footnotes

1 https://www.nature.com/articles/s41531-018-0058-0

## References

1- Alberico, S. L., Cassell, M. D., & Narayanan, N. S. (2015). The Vulnerable Ventral Tegmental Area in Parkinson’s Disease. Basal ganglia, 5(2-3), 51–55. https://doi.org/10.1016/j.baga.2015.06.001

2- Akandere, M., & Demir, B. (2011). The effect of dance over depression. Collegium antropologicum, 35(3), 651–656.

3- Bearss, K., & DeSouza, JFX. (2017). Plasticity in Damaged Multisensory Networks. In Thomas Heinbockel (Eds.), Synaptic Plasticity http://dx.doi.org/10.5772/67217:

4- Bearss, K., Mcdonald, K., Bar, R., & DeSouza, JFX. (2017). Improvements in balance and gait speed after a 12 week dance intervention for Parkinson’s disease. Advances in Integrative Medicine, 4(1), 10–13. doi:http://dx.doi.org/10.1016/j.aimed.2017.02.002

5- Berridge KC, Kringelbach ML.(2015). Pleasure systems in the brain. Neuron 2015; 86(3):646–664.

6- Bognar, S., DeFaria, A. M., O’Dwyer, C., Pankiw, E., Simic Bogler, J., Teixeira, S., Nyhof-Young, J., & Evans, C. (2017). More than just Dancing: experiences of people with Parkinson’s disease in a therapeutic dance program. Disability and rehabilitation, 39(11), 1073–1078. https://doi.org/10.1080/09638288.2016.1175037

7- Ciantar Sarah, DeSouza, J, Bears K. (2019). Investigating Affective and Motor Improvements with Dance in Parkinson’s Disease, BioRxiv (Preprint) Available at doi: 10.1101/665711 (Accessed Nov 30,2018).

8- Cummings, J. L. (1992). Depression and Parkinson’s disease: A review. The American Journal of Psychiatry, 149(4), 443– 454. https://doi.org/10.1176/ajp.149.4.443

9- DeSouza, JFX, & Bar, R. (2012). The effects of rehearsal on auditory cortex: An fMRI study of the putative neural mechanisms of dance therapy. Seeing and Perceiving, 25, 45–55.

10- DeSouza, J., & Bearss, K. (2018). Progression of Parkinson’s disease symptoms halted using dance over 3-years as assessed with MDS-UPDRS. Neuroscience Meeting Planner. San Diego: Society for Neuroscience Abstracts. Online, doi:https://abstractsonline.com/pp8/#!/4649/presentation/22209

11- Dils, Ann, and Albright, Ann Cooper, eds. (2001). Moving History/Dancing Cultures: A Dance History Reader. Middletown: Wesleyan University Press, ProQuest Ebook Central. (Accessed September 5, 2020).

12- Dorsey E.R., et al. (2018). Global, regional, and national burden of Parkinson’s disease, 1990– 2016: a systematic analysis for the Global Burden of Disease Study 2016, The Lancet Neurology, Volume 17, Issue 11, 2018, Pages 939–953, ISSN 1474-4422, https://doi.org/10.1016/S14744422(18)30295-3.

13- Dos Santos Delabary, M., Komeroski, I. G., Monteiro, E. P., Costa, R. R., & Haas, A. N. (2018). Effects of dance practice on functional mobility, motor symptoms and quality of life in people with Parkinson’s disease: a systematic review with meta-analysis. Aging clinical and experimental research, 30(7), 727–735. https://doi.org/10.1007/s40520-017-0836-2

14- Drew Daniel S, Muhammed Kinan, Baig Fahd, Kelly Mark, Saleh Youssuf, Nagaraja Sarangmat, Okai David, Hu Michele, Manohar Sanjay, Husain Masud (2020). Dopamine and reward hypersensitivity in Parkinson’s disease with impulse control disorder, Brain, Volume 143, Issue 8, August 2020, Pages 2502–2518, https://doi.org/10.1093/brain/awaa198

15- Fontanesi, C., & DeSouza, J. (2021). Beauty That Moves: Dance for Parkinson’s Effects on Affect, Self-Efficacy, Gait Symmetry and Dual Task Performance. Frontiers in Psychology,| https://doi.org/10.3389/fpsyg.2020.600440

16- Hackney E Madeline, Kantorovich Svetlana, Levin Rebecca, Earhart M Gammon. (2007). Effects of tango on functional mobility in Parkinson’s disease: a preliminary study. Journal of neurologic physical therapy, Vol 31(4), 173–179.

17- Hackney, M. E., and Earhart, G. M. (2009b). Health-related quality of life and alternative forms of exercise in Parkinson disease. Parkinsonism Relat. Disord. 15, 644–648. doi: 10.1016/j.parkreldis.2009.03.003

18- Hall, M. F. E., & Church, F. C. (2020). Exercise for Older Adults Improves the Quality of Life in Parkinson’s Disease and Potentially Enhances the Immune Response to COVID-19. Brain Sciences, 10(9) doi:10.3390/brainsci10090612

19- Harrar, V., Bar, R., & DeSouza, JFX. (2016). The auditory cortex changes across learning choreography with Parkinson’s Disease: fMRI changes across 8 months and a documentary. J Parkinsons Dis.;6 (supplement 1).

20- Hashimoto, H., Takabatake, S., Miyaguchi, H., Nakanishi, H., & Naitou, Y. (2015). Effects of dance on motor functions, cognitive functions, and mental symptoms of Parkinson’s disease: a quasi-randomized pilot trial. Complementary therapies in medicine, 23(2), 210–219. https://doi.org/10.1016/j.ctim.2015.01.010

21- Heiberger Lisa Et Al: (2011). Impact of a weekly dance class on the functional mobility and on the quality of life of individuals with Parkinson’s Disease, Cortical Motor Control Laboratory, Department of Neurology and Neurophysiology, University Hospital of Freiburg, Germany. Frontiers. Aging Neuroscience. | https://doi.org/10.3389/fnagi.2011.00014

22- Jankovic J. (2008). Parkinson’s Disease: Clinical features and diagnosis, Journal of Neurology, Neurosurgery & Psychiatry;79:368–376. http://dx.doi.org/10.1136/jnnp.2007.131045

23- Koch Sabine C, Morlinghaus Katharina, Fuchs Thomas. (2007).The joy dance: Specific effects of a single dance intervention on psychiatric patients with depression, The Arts in Psychotherapy, Volume 34, Issue 4, Pages 340–349, ISSN 0197-4556, https://doi.org/10.1016/j.aip.2007.07.001.

24- Levkov, G., Di Noto, P., Montefusco-Siegmund, R., Bar, R., & DeSouza, JFX. (2014). Global alpha slowing in individuals with Parkinson’s disease and dance-induced increases in frontal alpha synchronization. Neuroscience Meeting Planner Society for Neuroscience Abstracts, 437.

25- Pellicano, C., Benincasa, D., Pisani, V., Buttarelli, F. R., Giovannelli, M., & Pontieri, F. E. (2007). Prodromal non-motor symptoms of Parkinson’s disease. Neuropsychiatric disease and treatment, 3(1), 145–152. https://doi.org/10.2147/nedt.2007.3.1.145

26- Ng, T.H., Alloy, L.B. & Smith, D.V.(2019). Meta-analysis of reward processing in major depressive disorder reveals distinct abnormalities within the reward circuit. Transl Psychiatry 9, 293. https://doi.org/10.1038/s41398-019-0644-x

27- Quinn Lori, Chelsea Macpherson, Katrina Long, Hiral Shah.(2020). Promoting Physical Activity via Telehealth in People With Parkinson Disease: The Path Forward After the COVID-19 Pandemic?, Physical Therapy, Volume 100, Issue 10, October 2020, Pages 1730– 1736, https://doi.org/10.1093/ptj/pzaa128

28- Watson, D., & Clark, L. (1999). The PANAS-X: Manual for the positive and negative affect schedule - Expanded Form. Department of Psychological & Brain Sciences Publications, doi:10.17077/48vt-m4t2

29- Westheimer, O., McRae, C., Henchcliffe, C., Fesharaki, A., Glazman, S., & Ene, H., et al. (2015). Dance for PD: a preliminary investigation of effects on motor function and quality of life among persons with Parkinson’s disease (PD). Journal of Neural Transmission (vienna, Austria : 1996), 122(9), 1263–1270. doi:10.1007/s00702-015-1380-x.

30- Westheimer (2008). Why Dance for Parkinson’s Disease. Topics in Geriatric Rehabilitation, 24(2), 127–140.

31- https://danceforparkinsons.org/resources/dance-at-home

32- https://globalnews.ca/video/6844619/coronavirus-bringing-seniors-out-of-isolation-and-fostering-connection-through-free-online-dance-classes

33- https://www.movementdisorders.org/MDS/About/Movement-Disorder-Overviews/Parkinsons-Disease--Parkinsonism.htm

34- https://www.movementdisorders.org/MDS/About/Who-We-Are/Purpose-Mission--Goals.htm

